# Hepatitis C Core Antigen test as an alternative for diagnosing HCV infection: mathematical model and cost-effectiveness analysis

**DOI:** 10.1101/2019.12.12.19014621

**Authors:** Maryam Sadeghimehr, Olivia Keiser, Francesco Negro, Maia Butsashvili, Sonjelle Shilton, Irina Tskhomelidze, Maia Tsereteli, Barbara Bertisch, Janne Estill

## Abstract

**Background:** The cost and complexity of polymerase chain reaction (PCR) testing is a significant barrier for the diagnosis and treatment of patients infected with hepatitis C virus (HCV). We investigated the cost-effectiveness of various testing strategies using antigen as an alternative to PCR.

**Methods:** We developed a mathematical model for HCV to estimate the number of newly diagnosed individuals and cases of different stages of liver disease. We compared the following testing strategies: antibody test followed by PCR in case of positive antibody (baseline strategy); antibody test followed by HCV-antigen test (antibody-antigen); antigen test alone; and PCR test alone. We conducted cost-effectiveness analyses considering the costs of HCV testing (of both infected and uninfected individuals) (A1), liver-related complications (A2) and all costs including HCV treatment (A3). The model was parameterized for the country of Georgia, and several sensitivity analyses were conducted to generalize the findings for different settings.

**Results:** Using the current standard of testing, 89% of infected individuals were detected. Comparatively, antibody-antigen and antigen testing alone detected 86% and 88% of infected individuals, respectively. PCR testing alone detected 91% of the infected individuals with the remaining 9% dying or spontaneously recovering before testing. In analysis A1, antibody-antigen testing was not found to be essentially cheaper compared to the baseline strategy. In analysis A2, strategies using PCR were cheaper than antigen-based strategies. In analysis A3, antibody-antigen testing was the cheapest strategy, followed by the baseline strategy, and PCR testing alone.

**Conclusion:** Antigen testing, either following a positive antibody test or alone, performed almost as well as the current practice of HCV testing. The cost-effectiveness of these strategies strongly depends on the inclusion of treatment costs.

**Lay summary:** Core antigen testing is a reliable alternative test for diagnose HCV infection. Antigen-based strategies may be cost-effective, in particular if treatment costs are considered.

**Highlights:**

- Strategies using an antigen test to diagnose HCV infection performed reasonably well compared with the traditional antibody- and PCR based approach.
- According to our study, antigen test alone missed about 3%, and antibody followed by PCR test 2% of HCV infected individuals.
- The maximum difference in quality-adjusted life expectancy across the different strategies of diagnosing HCV was only one month.

## Introduction

Hepatitis C virus (HCV) is a major cause of liver disease and liver-related mortality.^1-3^ The World Health Organization (WHO) estimates that approximately 71 million people worldwide are chronically infected with hepatitis C, and almost 400,000 people die from HCV every year, mostly due to cirrhosis and hepatocellular carcinoma. However, the majority of the HCV infected individuals are not aware of their infection.^4^ Simple and effective hepatitis testing strategies and tools are needed to achieve the WHO target of eliminating HCV as a major public health threat by 2030.^5^

Since 2014, Direct Acting Antivirals (DAA) form the standard treatment for HCV. To be successfully treated with DAA, diagnostic tests are needed to diagnose the infected patients and confirm the clearance of viral replication 12-24 weeks after the end of treatment.^6-8^ Two types of tests are usually used for diagnosing HCV: serological assays that detect antibodies to HCV, and nucleic acid tests that detect HCV RNA genomes in order to confirm active infection.^9^ The most commonly used testing protocol is to first use an antibody test, and in the case of a positive test result, to use a nucleic acid test (most commonly a polymerase chain reaction test, PCR) to check the presence of the virus.^6^ The sensitivity and specificity of PCR tests are high.^9^ PCR testing requires time and trained laboratory personnel, which increases the costs. The cost of PCR testing is an important barrier in implementing comprehensive testing, especially in low- and middle-income countries.

HCV antigen testing is a serological assay that directly detects a viral protein and can give a positive test result as soon as the virus component is present. This test can be done on the same platform as the antibody test.^8^ Antigen testing is cheaper^10^ and requires less special training than PCR testing. While antigen tests have a specificity of up to 100%, viral loads below 3000 IU/ml may not be detected.^7,11-13^

With a limited budget, replacing PCR by antigen testing could increase testing coverage, but people with very low viral loads may be missed. Using the country of Georgia as an example, we aimed to study the cost-effectiveness of different testing strategies using a mathematical model.

## Materials and methods

### Model structure and inputs

We developed a mathematical model for HCV disease progression, similar to a previously published model.^14^ We simulated cohorts of patients who were followed from the time of infection until death. The progression of HCV is represented by a directed acyclic graph of health states. In each state, the model samples transition times to all possible destination states. The minimum of these times determines when and to which state the patient will move next. The process is repeated until the patient reaches a terminal state (death). The patients progress along two dimensions: progression of liver disease, and course of the HCV infection including cascade of care (Figure 1). The condition of liver disease is represented by the METAVIR scoring system (F0 to F4), followed by decompensated cirrhosis (DC), hepatocellular carcinoma (HCC) and liver transplantation (LT). Death is represented using separate states depending on the cause of death: liver-related death, HIV-related death, drug use related death, and death due to other causes. At the beginning of the simulation, patients are assigned the following characteristics: age at infection, year of birth, gender, HIV co-infection, level of alcohol consumption, and duration of intravenous drug use (IDU).

**Figure 1.**
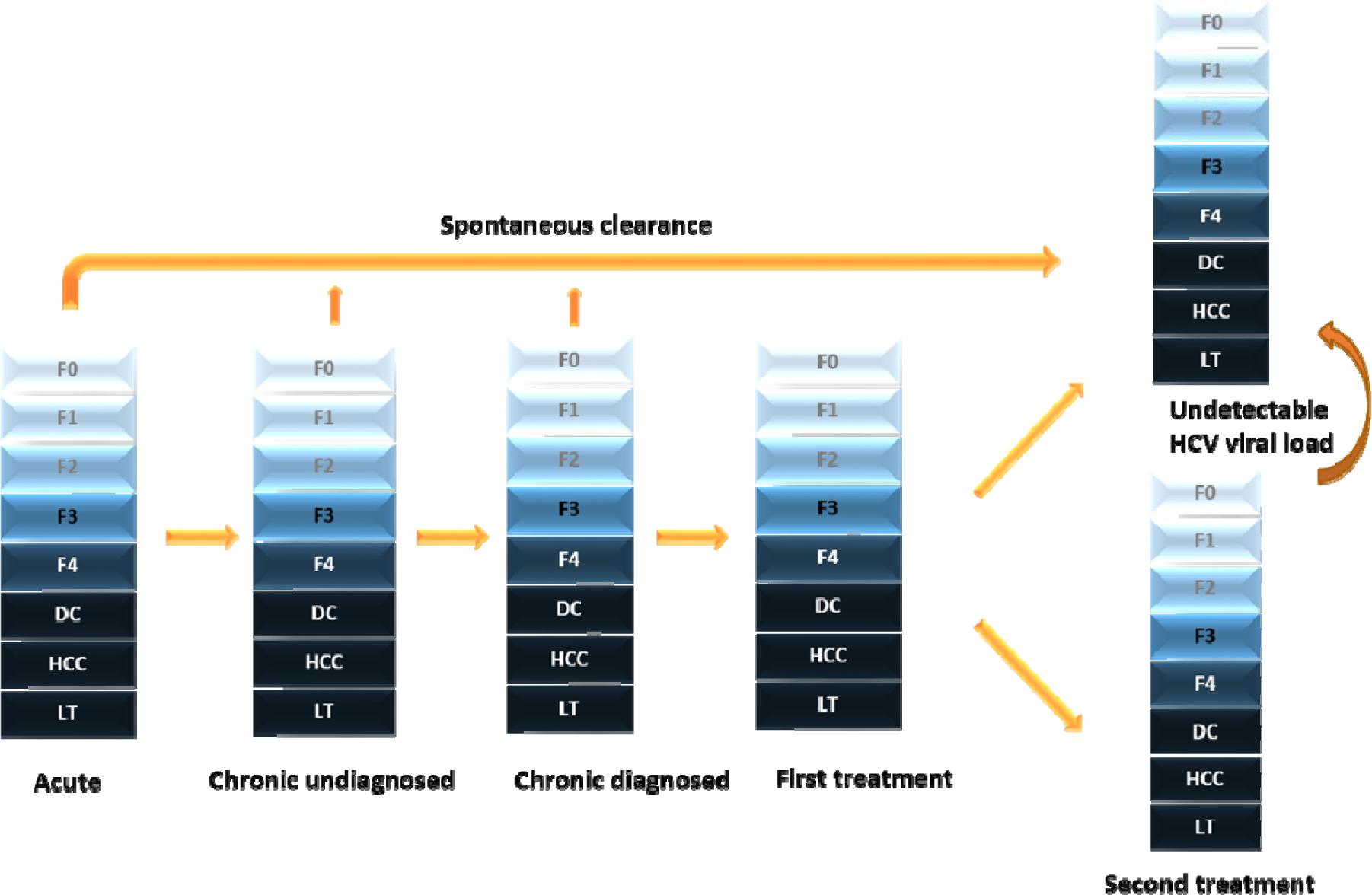
Structure of the simulation model. Individuals can progress vertically based on liver disease, and horizontally through the hepatitis C virus (HCV) infection and cascade of care. First and second treatments contain the treatment episode itself and, in case of treatment failure, the time after ending therapy. Death can occur at any state (not shown in the graph for simplicity). F0-F4: stages of fibrosis according to the METAVIR scoring system; DC: decompensated cirrhosis; HCC: hepatocellular carcinoma; LT: Liver transplantation.

Many studies have shown that hepatitis C viral load is relatively stable in untreated patients with chronic infection.^9,15-16^ We therefore assumed that viral load remains approximately constant in untreated individuals. We used the distribution of hepatitis C viral load among patients in the Swiss Hepatitis C Cohort Study (SCCS) database^11^ and assigned each patient a baseline viral load. Viral load values at the time of HCV testing were sampled from a log-normal distribution around the baseline viral load. We denote viral loads below 3000 IU/ml as very low viral loads (VLVL). 7,11-12

We considered the following strategies of testing: HCV-antibody followed by PCR testing in case of a positive antibody test (baseline strategy); HCV-antibody followed by HCV-antigen testing in case of a positive antibody test; HCV-antigen test alone; and PCR test alone. In all strategies, a second test (either PCR or antigen, whichever was used for confirming the diagnosis) was taken 12 weeks after completion of treatment to confirm sustained virologic response (SVR). We assumed that all individuals were tested for HCV once during the years 2015 to 2018. In case of a negative test result, the individual was not retested.

The probability of HCV detection depends on the rate of HCV testing and the sensitivity of the test. We assumed that the probability of HCV detection by antibody test increases exponentially during the first year of the infection and stabilizes at 99% thereafter. The sensitivity of the antigen test was assumed to be 98.2% for patients with a viral load above 3000 IU/ml, and 33.0% for patients with VLVL.^11^ The PCR test was assumed to be 100% sensitive. As our model only simulates infected patients, the expected numbers of tests among HCV uninfected people were calculated from the estimated HCV prevalence in the target population. We assumed 100% specificity for HCV-antigen and PCR tests.

We assumed that all detected patients are treated with DAAs, and 98% of the treated patients achieve SVR^6^. The parameters related to liver disease progression are shown in <u>Supplementary Table 1</u>. We assumed that HCV viral load was not independently associated with fibrosis progression rates.^2, 17^

We parameterized the model to represent the epidemic in the country of Georgia. Georgia was among the first countries that aimed to eliminate HCV and started an HCV elimination program^18^ in April 2015. Enlarged-scale HCV screening began already in January 2015, before the launch of the program. Screening services continue to be provided in various settings free of charge. As of June 30, 2018, a total of 1,175,291 HCV screening tests had been done and 1,125,808 persons registered in the elimination program, of whom 93,181 (8.3%) were positive for HCV antibody. Among persons receiving HCV testing, most were screened during inpatient hospitalization (44%); other groups of persons receiving HCV screening included blood donors (18%), and pregnant women (9%).^19^ The current laboratory testing process in Georgia is as follows: patients with unknown or no documented HCV serological status first undergo anti-HCV antibody testing by rapid or laboratory-based methods (i.e., enzyme-linked immunosorbent assay [ELISA] or chemiluminescent immunoassay [CIA]). Patients with documented HCV serological status and positive anti-HCV antibodies undergo testing to determine active HCV infection by PCR, or since December 2017 alternatively with core antigen testing.^20^ We assumed that HCV prevalence was 66.2% among IDUs and 5.4% among the rest of the population.^21-22^ IDUs represented 25% of the simulated cohort.^19^ Table 1 and <u>Supplementary Figures 1-2</u> present the detailed baseline characteristics of the simulated individuals. The simulated population included only patients who were infected before the year 2019, and who had not cleared the virus spontaneously or been treated before 2015. We only considered the infected individuals who were not diagnosed by the year 2015.

**Table 1.** Baseline characteristics of the simulated patients. IDU, injection drug user; MSM, men having sex with men; DC, decompensated cirrhosis; HCC, hepatocellular carcinoma; F3, F4: fibrosis stage according to METAVIR score.

| Baseline characteristics of the simulated patients |  |  |  |
| --- | --- | --- | --- |
| Characteristics | active IDU | non-IDU | Source |
| Alcohol consumption |  |  |  |
| Abstinent | 37% | 37% | 36, assumption |
| Moderate (on average 20-40 g per day) | 37% | 37% |  |
| Excessive (on average > 40 g/d) | 26% | 26% |  |
| Gender |  |  |  |
| Female | 0.8% | 47.2% | 36, 37, 18 |
| Male | 99.2% | 52.8% |  |
| HIV co-infected | 2.3% | 0.2% | 38 |
| HCV prevalence | 66.2% | 5.4% | 20, 39, 40 |

### Model outcomes

We estimated the number of diagnoses for each testing strategy and compared the number of people who experienced severe liver disease (F3), cirrhosis (F4), DC, HCC, and liver-related death within their lifetime.

We also conducted cost-effectiveness analyses comparing the different testing alternatives. We conducted three analyses with different assumptions regarding costs. In Analysis A1, we only considered the direct costs of the HCV tests (diagnosis and treatment monitoring), including also HCV uninfected individuals who were not explicitly simulated. In Analysis A2, we included in addition other laboratory costs during treatment (including clinical assessment, complete blood count, HCV-associated consultations, and alanine aminotransferase (ALT) test)^20^ as well as the lifetime costs associated with liver disease. This analysis takes the perspective of the health care payer in situations like in the country of Georgia where treatment costs are covered by external donors. In Analysis A3, we included all costs of HCV testing, liver disease and HCV treatment. We reviewed the literature and contacted persons directly involved in the elimination project in the country of Georgia who helped to interpret already published data to obtain costs related to HCV testing, DAA treatment and liver disease, as well as HCV- and liver-related utilities (Table 2).21-24 As studies reporting average costs related to liver disease are scarce, we decided to adopt the life-time costs from Turkey for viremic individuals, and modified the costs in a sensitivity analysis.^23^ We assumed that the costs of liver disease in stages F0 to F3 decreased by 50% after achieving SVR. In all analyses, we calculated the incremental cost-effectiveness ratios (ICERs) between the strategies, comparing the incremental costs with incremental gain in quality-adjusted life expectancy at time of infection.

**Table 2.** Unit costs and health utilities.

| Costs | Value | Source |
| --- | --- | --- |
| Antibody test | \$2 | 20, 41 |
| Antigen test | \$21 | |
| PCR test | \$40 | |
| Physician visit and blood collection | \$13 | |
| Treatment monitoring costs<sup>1</sup> | \$117 | 20 |
| Treatment cost | \$ 50,674 | 42,43 |
| Average annual cost of liver disease |  |  |
| Fibrosis stage F0-F2 | \$447 | 44 |
| Fibrosis stage F3 | \$447 | |
| Fibrosis stage F4 | \$578 | |
| Decompensated cirrhosis | \$1984 | |
| Hepatocellular carcinoma | \$2474 | |
| Health-related utilities: |  |  |
| Fibrosis stage F0-F2 | 0.82 | 23, 24 |
| Fibrosis stage F3 | 0.76 |  |
| Fibrosis stage F4 | 0.76 |  |
| Decompensated cirrhosis | 0.60 |  |
| HCC | 0.60 |  |
| F0-F1 after sustained virological response | 0.95 |  |
| F2-F4 after sustained virological response | 0.85 |  |

<sup>1</sup> Including the cost of clinical assessment, complete blood count, ALT (AST, creatinine), patient service standard

The results are presented per infected individual. We discounted all future costs and quality-adjusted life years (QALYs) at 3% per year.

### Sensitivity analyses

In order to address the uncertainty around key parameters and generalize our findings to other settings, we conducted several sensitivity analyses (<u>Supplementary Table 2</u>). First, we modified the costs of tests by reducing the unit cost of either the PCR test (sensitivity analysis S1) or the antigen test (sensitivity analysis S2). Second, we reduced the liver-related costs after SVR to zero for liver stages F0-F2 (sensitivity analysis S3). Third, we used an alternative estimate of costs for liver diseases from France (sensitivity analysis S4). Finally, we calculated the results for a population consisting completely of non-IDUs with decreased HCV prevalence (sensitivity analysis S5), or an IDU population with an increased HCV prevalence (sensitivity analysis S6).

## Results

In the baseline scenario, 89,400 persons out of 100,000 infected individuals were diagnosed during the four-year screening period. In the strategy with antibody followed by antigen testing, fewer infected individuals were detected (86,100 persons per 100,000 infected individuals). For antigen test alone the number of diagnoses was 87,500 persons per 100,000 infected individuals. PCR test alone could detect the highest number of infected individuals (91,000 persons): the remaining 9% of infected individuals either died or spontaneously recovered before testing.

The proportion of patients who experienced severe liver disease was highest for antibody testing followed by antigen testing, and lowest for PCR testing alone (Figure 2). In the baseline strategy, 22.5% of infected individuals experienced at least liver disease stage F3 in their life. The percentages of people who experienced at least liver disease stage F3 were 23.5%, 22.9%, and 21.7% for antibody followed by antigen test, antigen test alone, and PCR test alone, respectively. The percentages of people who reached stage F4 ranged between 10.0% and 12.0% across all strategies. For DC, HCC, LT and liver-related death the corresponding ranges were 2.2%-3.0%, 1.4%-2.1%, 0.5%-0.6% and 3.7%-5.0%, respectively.

**Figure 2.**
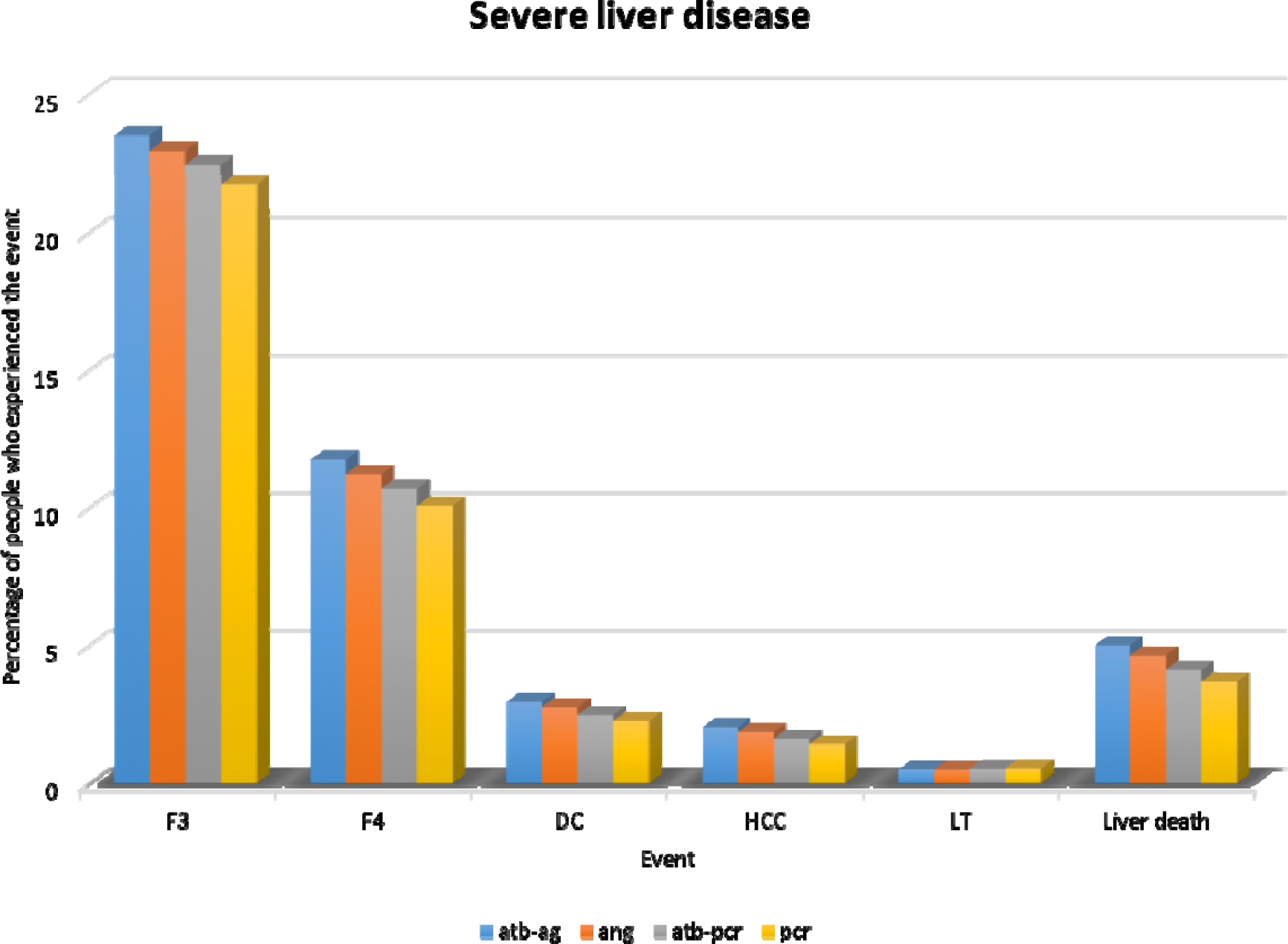
A comparison between different testing strategies: The proportion of infected individuals who experienced different stages of liver disease in their lifetime. DC: decompensated cirrhosis; HCC: hepatocellular carcinoma; LT: liver transplantation; atb: antibody; ang: antigen.

Figure 3A presents the cost versus QALY for Analysis A1, considering only the cost of testing. Antibody followed by antigen testing was the cheapest strategy with a total cost of $215 per infected individual a mean quality-adjusted life expectancy of 15.51 QALYs. The most cost-effective strategy compared with antibody followed by antigen was the baseline strategy, with a quality-adjusted life expectancy of 15.56 QALYs and an ICER of $369/QALY gained. Antigen alone had higher costs than the baseline strategy. PCR alone, which had a mean quality-adjusted life expectancy of 15.60 QALYs, was the most effective strategy with an ICER of $10,763/QALY gained compared with the baseline strategy.

**Figure 3.**
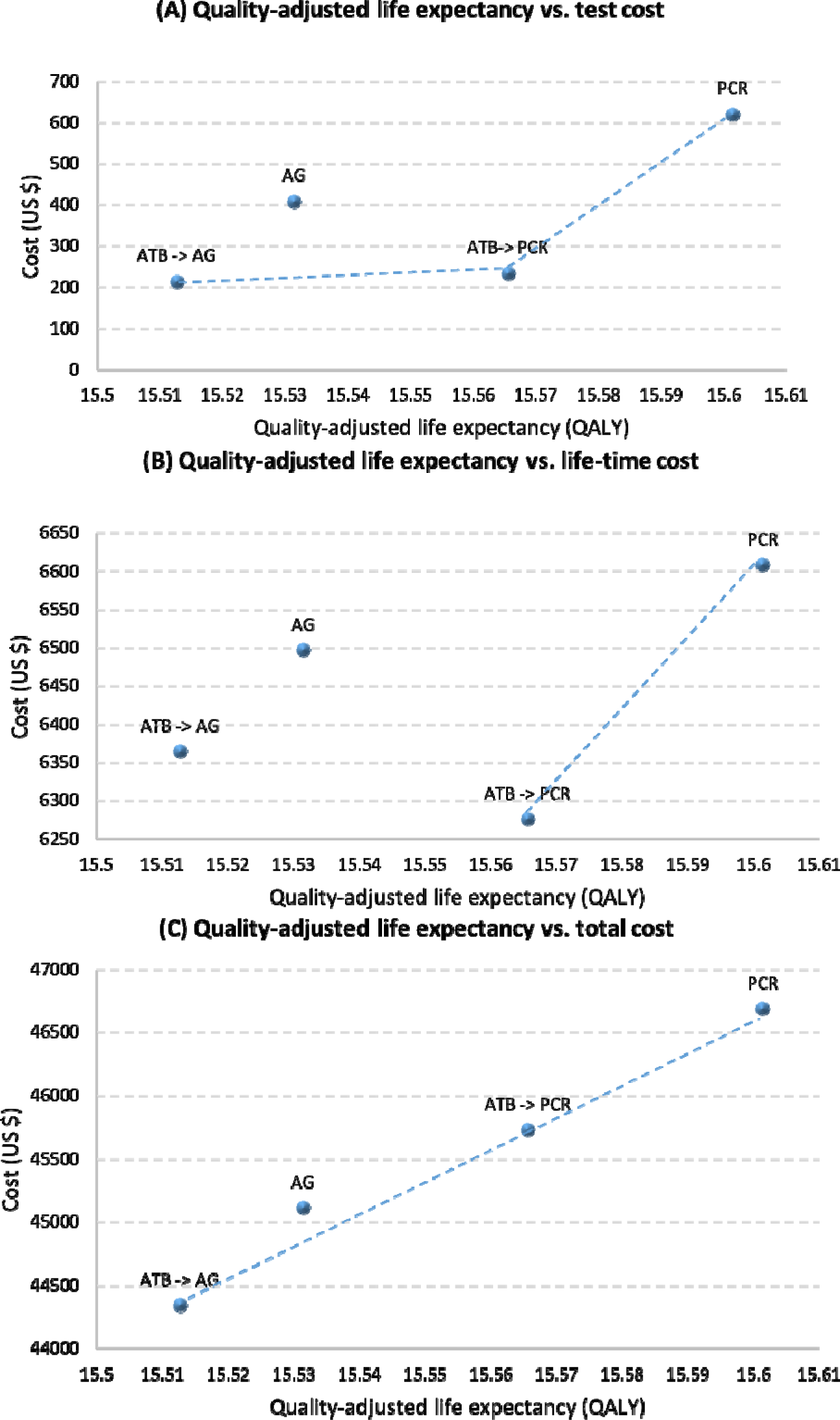
Quality-adjusted life expectancy versus cost. (A) Analysis 1: Cost of HCV testing versus the quality-adjusted life expectancy. (B) Analysis 2: Cost of HCV testing and life-time liver-related complications versus the quality-adjusted life expectancy. (C) Analysis 3: Cost of HCV testing, life-time liver related complications and HCV treatment versus the quality-adjusted life expectancy. All costs are measured per infected individual and include also costs of negative tests. Quality-adjusted life expectancy is measured per infected individual at the time of infection. All QALYs and costs are discounted by 3% per year. QALY: quality-adjusted life years; ATB: antibody; AG: antigen.

Life-time liver disease costs including the cost of laboratory testing and treatment monitoring (Analysis A2) are presented in Figure 3B. In this analysis, the baseline scenario was the cheapest scenario with a life-time cost of $6,275. PCR test alone, the only strategy performing better than the baseline, had an ICER of $9,281/QALY gained compared with the baseline scenario.

In Analysis A3 considering all costs of testing, liver disease and treatment, antibody followed by antigen was again the cheapest scenario, with an average life-time cost of $44,340 (Figure 3C). Compared with antibody followed by antigen testing, the baseline strategy was the most cost-effective strategy with an ICER of $26,363/QALY gained. Compared with the baseline, the ICER of PCR testing alone was $26,643/QALY gained.

### Sensitivity analyses

Changing the input costs of diagnostic tests or liver disease did not change the patterns of cost-effectiveness substantially (<u>Supplementary Figures 3-6</u>). The largest differences were in the analyses of the low- and high-prevalence populations. In the case of a low-prevalence non-IDU population, the results of all three analyses were driven by the costs of testing the negative individuals. Replacing the two-step testing (i.e. antibody test followed by PCR or antigen test) with antigen alone increased the costs of testing (analysis A1) by $2,000 per infected individual; for PCR alone costs increased by $4,000 per infected individual (<u>Supplementary Figure 7</u>). In the high HCV prevalence IDU population, the situation was reversed (<u>Supplementary Figure 8</u>). Considering the costs of testing only (analysis A1), antigen and PCR testing alone were slightly cheaper than their corresponding two-step procedures. If costs of liver disease were also included (analysis A2), PCR testing alone was even the cheapest strategy. Considering all costs (analysis A3), antibody followed by antigen was again the cheapest scenario. Of the remaining strategies, antigen testing alone was the most cost-effective strategy with an ICER of $12,265/QALY gained compared to the cheapest strategy.

## Discussion

### Principal findings

Strategies using an antigen test to diagnose HCV infection performed reasonably well compared with the traditional PCR based approach, but the cost-effectiveness of these strategies depends on the perspective taken. In situations like in the country of Georgia, where treatment is provided from external sources^20^, the current two-step testing procedure using antibody and PCR tests has the lowest costs from the healthcare system’s point of view. When we added the HCV treatment costs to our analysis, the two-step procedure with antigen instead of PCR as the confirmatory test was the cheapest, but also the least effective, strategy. The maximum difference in quality-adjusted life expectancy across all strategies was however only one month. Antigen testing alone performed better than antibody followed by antigen, but not as well as the baseline strategy. PCR testing alone was clearly the most effective but also most expensive strategy. These additional costs could however be compensated by cost savings related to liver disease, if we did not consider the costs of HCV treatment.

Antigen testing alone has been considered a potential alternative for the current two-step testing procedure, which requires the patient to visit the clinic multiple times. Both strategies miss some HCV infected individuals. According to our study, antigen alone missed about 3% and the baseline strategy 2% of those infected. The characteristics of the missed patients are however different. The HCV antibody test can detect a positive result only about 35 days after infection^25^, and strategies using antibody tests as the initial test, such as the baseline strategy, may lead to possible under-diagnosis in populations with ongoing transmission. However, the Georgian HCV epidemic can be characterized as an “old epidemic”: most HCV infections were acquired during the first years after the collapse of the Soviet Union.^26^ Also, in immunosuppressed patients, the antibody test may always be negative.^27^ The antigen test in turn misses around two thirds of individuals with VLVL. Antigen testing was less beneficial than the baseline strategy for two reasons: the number of VLVL patients was higher in patients with chronic infection than in recently infected patients, and VLVL patients, as chronically infected persons, reach end-stage liver disease on average earlier than those recently infected. Antigen testing saved costs mainly by missing the VLVL infected individuals and therefore reducing the number of treated patients. However, spontaneous cure is frequent among VLVL individuals, as compared to chronically infected patients^11^. Moreover, if low viral load values would slow down the progression of liver disease, antigen might no longer be inferior to the baseline. The probability of onward HCV transmission may also be clearly lower in persons with VLVL.^28^ Using a one-step simple test could have other advantages, such as reducing the risk of loss to follow-up (LTFU). In Georgia, more than 25% of anti-HCV positive individuals did not get a confirmatory test and were considered LTFU in 2015.^18^ Using a one-step simple test would also reduce the unnecessary anxiety among individuals with a false positive result or with spontaneous cure.

Although PCR testing is more expensive than antigen testing, our analysis revealed some situations where the total costs may be lower with PCR based strategies. If the costs of treatment are not considered, the costs saved by preventing progression to advanced liver disease in a few patients could outnumber the additional costs needed for testing by PCR, either according to the current practice, or even by using PCR alone. In addition to the country of Georgia, where treatment is covered by external sources, this may also be relevant for other countries that have negotiated special agreements with treatment manufacturers. In the “subscription model”^29,30^, where the government pays a flat fee for treating all infected residents within a given time period, the costs of treatment do not depend on the number of treated patients, and the cost-effectiveness evaluations should thus focus on the costs of liver disease and diagnostics. PCR testing alone was used in our analysis as a theoretical best-case comparator: testing the population with this test costing more than $40 is unlikely. However, in a setting with extremely high prevalence and ongoing transmission, such as active IDU, PCR testing alone could be cost-saving.

In a setting with a limited health budget, investments in less expensive tests can free resources to test and treat more infected individuals. When comparing the current practice to the less expensive strategy using antibody followed by antigen testing, the ICER was about $19,600 per QALY if all costs including treatment were included, and the high costs were mainly caused by the increased need of treatment. This shows that in higher-income countries, the use of PCR as the confirmatory test can be justified from the financial point of view. However, in low-income settings, it may not be efficient to invest in PCR tests, in particular if those with low viral loads have a lower rate of HCV transmission and high probability of spontaneously recovery. The formal cost-effectiveness analysis also does not take into account other factors such as budget restrictions. If only a fixed budget is available for testing within a given time period, the use of less expensive tests allows to test more individuals, thus leading to more diagnoses and better clinical outcomes. Under some conditions, the use of antigen as a confirmatory test may thus be a beneficial solution.

### Strengths and limitations

Several studies have compared antigen and PCR testing in different settings and proposed the use of antigen testing.^24,31,33-34^ These studies were however usually limited to compare the costs of the diagnosis, or other short-term costs, alone. Our study compares different HCV testing strategies on the life-time burden of HCV infection in a nationwide setting.

Our study is subject to limitations. First, HCV transmission was not included. We ignored the additional burden of the disease and costs that each missed case might cause due to onward transmission of HCV. A massive scale-up of therapy may reduce the number of new infections. Second, we did not consider HCV reinfection after achieving SVR. Reinfection has been might reduce the overall difference in QALYs as well as increase the overall costs. Third, we did not model any extrahepatic manifestations (EHM) explicitly. EHM could increase the overall life-time costs associated with HCV.^35^ Including the costs of EHM could further favor the more effective strategies. Fourth, we did not allow for HCV re-testing. Individuals who identify themselves at high risk of infection, such as active IDUs, are recommended to get retested at least once a year.^6^ This is also the population with most acute infections. As antibody testing misses those recently infected, our model may overestimate the benefit of strategies with antibody testing for populations with a high number of acute infections. Fifth, we did not take into account some factors that could favor the use of antigen testing, such as the possibly slower progression, lower risk of onward transmission and higher rate of spontaneous cure among patients with VLVL, the potentially better retention of one-step testing strategies, and potential reductions of the cost of antigen testing. Sixth, our analyses took the perspective of a health care payer. Although in the Georgian elimination program all costs are now paid from the governmental expenditure, in other settings where a considerable part of the costs is covered with out-of-pocket payments the question of finding the most cost-effective strategy becomes more complicated.

## Conclusions

A single antigen test can be a reliable and practical alternative for the current two-step procedure to diagnose HCV infection, but the cost-effectiveness of this strategy depends on various factors. In settings where the costs of treatment do not directly depend on the number of treated patients, the higher costs of PCR testing are likely to be compensated by the savings in liver disease related costs. However, a full consideration of treatment related costs may favor simpler and easier tests such as antigen alone, or antigen after antibody testing. The change to a simple one-time test could offer several advantages, and the vast majority of individuals chronically infected with HCV have viral loads above 3000 IU/ml, a level at which the diagnostic capacity of antigen tests does not differ from PCR. The policy-makers should therefore carefully review the situation in each country to decide the optimal strategy of diagnosing HCV infection.

## Data Availability

We reviewed the literature and contacted persons directly involved in the elimination project in the country of Georgia who helped to interpret already published data.

## Acknowledgements

This study was funded by Gilead Switzerland Sarl. OK was funded by a professorship grant from the Swiss National Science Foundation (no 163878). We would like to thank all the collaborators and study nurses of the Swiss Hepatitis C Cohort Study; and our colleague Matteo Brezzi, who recently passed away unexpectedly, for assisting with data collection and management.

